# Comparison Of Low-Density Lipoprotein Cholesterol (Ldl-C), Atherogenic Index In Plasma (Aip) And Apolipoprotein B/Apolipoprotein A1 Ratio In Patients Sent For Routine Lipid Profile Testing At Biochemistry Laboratory - Kenyatta National Hospital

**DOI:** 10.1101/2023.10.25.23297527

**Authors:** Peter Kibet Kiptim, Rahab Wangari Kariuki, Damaris Kimonge

**Affiliations:** Kenyatta National hospital, Department of laboratory medicine; Kenyatta National hospital, Department of medical research and programs

**Keywords:** Cardiovascular disease risk, Lipid profile, Atherogenic index of plasma, Apo B, Apo A1, LDL-C

## Abstract

**Background:** Cardiovascular disease is a rising global concern accounting for one-third of mortalities. Notably, cardiovascular disease risk factors; including dyslipidemia, smoking, obesity, poor diet, and hypertension are modifiable. It has been established through various studies that dyslipidemia is at the core of atherosclerosis observed in cardiovascular diseases making it important in risk assessment and as a therapeutic target as informed by various guidelines. Plasma lipid profile has been key in the diagnosis of dyslipidemia particularly low-density lipoprotein cholesterol that is used in initial risk assessment and treatment monitoring. However, due to clinical and analytical challenges, LDLC is understated or overstated in some patients due to biological variability.

**Objective:** To compare low-density lipoprotein cholesterol, atherogenic index in plasma, and Apolipoprotein A1 / Apolipoprotein B ratio for cardiovascular risk assessment in patients sent for routine lipid profile testing at biochemistry laboratory - Kenyatta National Hospital

**Methodology:** This was laboratory-based cross-sectional study, 307 residual samples from patients sent for lipid profile testing were tested for, Apo A1, and Apo B concentrations using Mindray BS 2000M chemistry analyzer using Mindray reagents (Shenzhen Mindray Bio-Medical Electronics Co., Ltd) at biochemistry laboratory KNH. The obtained data was used to calculate the atherogenic index of plasma (AIP) and Apo B/ Apo A ratio.

**Results:** The median age of the participants was 49 years, 56% were female. The median(range) values were Total cholesterol 4.17 (3.11, 5.20),High-density lipoprotein cholesterol 1.21 (0.88, 1.54)Low-density lipoprotein cholesterol 2.60 (1.84, 3.37)Triglycerides 1.12 (0.83, 1.62),APOA-1 1.38 (1.11, 1.57),APOB 0.95 (0.73, 1.21)Apo B/ApoA-1 ratio 0.75 (0.56, 0.94) and Atherogenic index of plasma 0.09 (−0.14, 0.45). More than a quarter (26.7%) of the participants had LDL-C above 2.35 mmol/l. More than a third of the patients (42%, n=124) were at risk of CVD according to ApoB/ApoA-1 ratio, while 40 %,(n=122) had a high CVD risk according to the atherogenic index stratification.

We observed a positive correlation between LDL-C and ApoA-1, Apo B and Apo B/ApoA-1 ratio. There is a significant agreement between atherogenic index and APOB/APOA-1 ratio in CVD risk identification kW=0.36 (95% CI, 0.27-0.46), p<0.001.

**Conclusion:** Atherogenic Index of plasma, Apo A and Apo B markers offer additional benefits in CVD risk identification.

## INTRODUCTION

The World Health Organization defines cardiovascular disease (CVD) as a catch-all term for a wide range of pathologies affecting the cardiovascular system. These include coronary heart disease (CHD), cerebrovascular disease, peripheral arterial disease, rheumatic heart disease, deep vein thrombosis, and pulmonary embolism (1). CVD represents one-third of deaths worldwide, with low and middle-income countries accounting for three-quarters of CVD deaths (1). Paradoxically, the most significant CVD risk factors, such as dyslipidemia, hypertension, obesity, physical inactivity, poor diet, and smoking, are modifiable. In 2010, the American Heart Association issued guidance for the population to lessen the burden of cardiovascular disease (CVD) by achieving seven ideal cardiovascular health (CVH) behaviors and factors known as “Life’s Simple 7”, which addressed smoking, nutrition, physical activity, body mass index, blood pressure, total cholesterol, and fasting glucose (2). Globalization, coupled with few measures and policies in place to preclude or control cardiovascular risk factors, has led in the rise in CVD particularly in the developing countries. (3–5).

Dyslipidemia is a modifiable risk factor, and it is intimately linked to CVD, making it a target for risk assessment, therapy, and monitoring (6). Typical dyslipidemia is characterized by an increase in plasma concentrations of low-density lipoprotein cholesterol (LDL-C), total cholesterol, and triglycerides with a decrease in plasma concentrations of high-density lipoprotein cholesterol (HDL-C) (4, 6). Numerous studies have revealed that LDL-C is an essential driving force in the advancement of atherosclerotic cardiovascular disease. The analysis of LDL cholesterol (LDLC) is a key aspect in assessing for cardiovascular disease (CVD) and monitoring for dyslipidemia (7, 8). The importance of LDL particles in the pathological process of atherosclerotic CVD cannot be overstated (8). Furthermore, randomized controlled trials and meta-analyses have discovered an explicit weighted link between LDL-C concentration and CVD (7, 10–11). However, with the upsurge of insulin resistance, metabolic syndrome, and long-term statin use, there is upsurge in patients with atherogenic dyslipidemia who have "normal" total cholesterol and LDL-C concentrations (12).LDL-C quantification is impacted by analytical challenges and biological variability reducing its reliability in predicting cardiovascular risk(7,12). Apolipoprotein B (Apo B) a structural protein component present in all non-HDL lipoproteins and can be used to quantify the concentration atherogenic lipoproteins in circulation (LDL, remnant lipoproteins, and Lipoprotein (a) (7, 13). Apo B stands out because it is more accurate than direct LDL-C and HDL-C assays and is unaffected by biological variation in triglycerides regardless of fasting state (7, 15). In the Copenhagen Heart Study, Apo B demonstrated a higher relative risk of CVD than LDL-C. (15). Another study discovered that including Apo B in addition to standard risk factors improved long-term CVD risk assessment (16).

In this study, we aimed to determine the clinical utility of Apo B, in cardiovascular disease risk assessment by comparing Apo B/Apo A1, LDL -C and atherogenic index of plasma (AIP) from remnant samples of patients who had been sent to for lipid profile testing at the biochemistry laboratory in Kenyatta National Hospital, Nairobi Kenya.

## METHODS

This was a laboratory based cross-sectional study conducted at Kenyatta National Hospital, department of laboratory medicine, Biochemistry laboratory. All samples that were submitted to biochemistry laboratory from consenting clients for routine lipid profile testing in the months of December 2022 and January 2023 were eligible to be included in the study. Consecutive sampling was employed until the sample size was achieved. The blood samples were allowed to stand for 30 minutes to allow the blood to clot before being spun at 4000 rpm for 5 minutes. The serum was then be aspirated and transferred to a secondary sample cup labeled appropriately and analyzed for requested lipid profile (TC, TG, HDL-C, LDL-C), Apo A1, Apo B using the Mindray BS 2000M analyzer using Mindray reagents(Shenzhen Mindray Bio-Medical Electronics Co., Ltd). Ethical approval was granted from the institutional research and ethics committee (KNH-UoN ERC). Approval number P553/06/2022

### Data Analysis

Microsoft Excel 2016 was used to enter raw data before it was transferred to STATA version 13 (Stata Corp Inc., USA) for analysis.

Descriptive statistics was used for continuous variables into frequencies and proportions to stratify CVD as low risk using AIP was computed Log of Triglyceride /HDL-C cut off of 0.11, intermediate risk values of 0.11-0.21 and increased risk using values greater than 0.21. (17) Descriptive statistics was used for continuous variables into frequencies and proportions to stratify CVD as low risk using Apo A1/Apo B ratio with cut off > 0.8 to classify those at risk. (18)Correlation analysis was used to compare the relationship between Apo A1, Apo B to LDL-C.

Kappa Statistic was used to determine level of agreement between ApoA1/Apo B ratio and atherogenic index of plasma in risk assessment of cardiovascular disease (CVD).

A kappa score of more than 0.81 meant an almost perfect agreement, 0.61-0.80 as substantial agreement, 0.41-0.60 as moderate agreement and 0.21-0.40 as a fair agreement.

## RESULTS

The median age of the study participants was 49 (38, 62) with a majority (56%, n=172) being females. The median value of the total cholesterol was 4.17 (3.11, 5.20). The median values of APOA-1 and APO B were 1.38 (1.11, 1.57) and 0.95 (0.73, 1.21) respectively. Twenty-two percent (n=66) of the patients had low values of APOA-1 and fifteen percent (n=45) had low values of APO B. The median value of low-density lipoprotein cholesterol was 2.60 (1.84, 3.37) as shown in Table1.

**Table 1:**
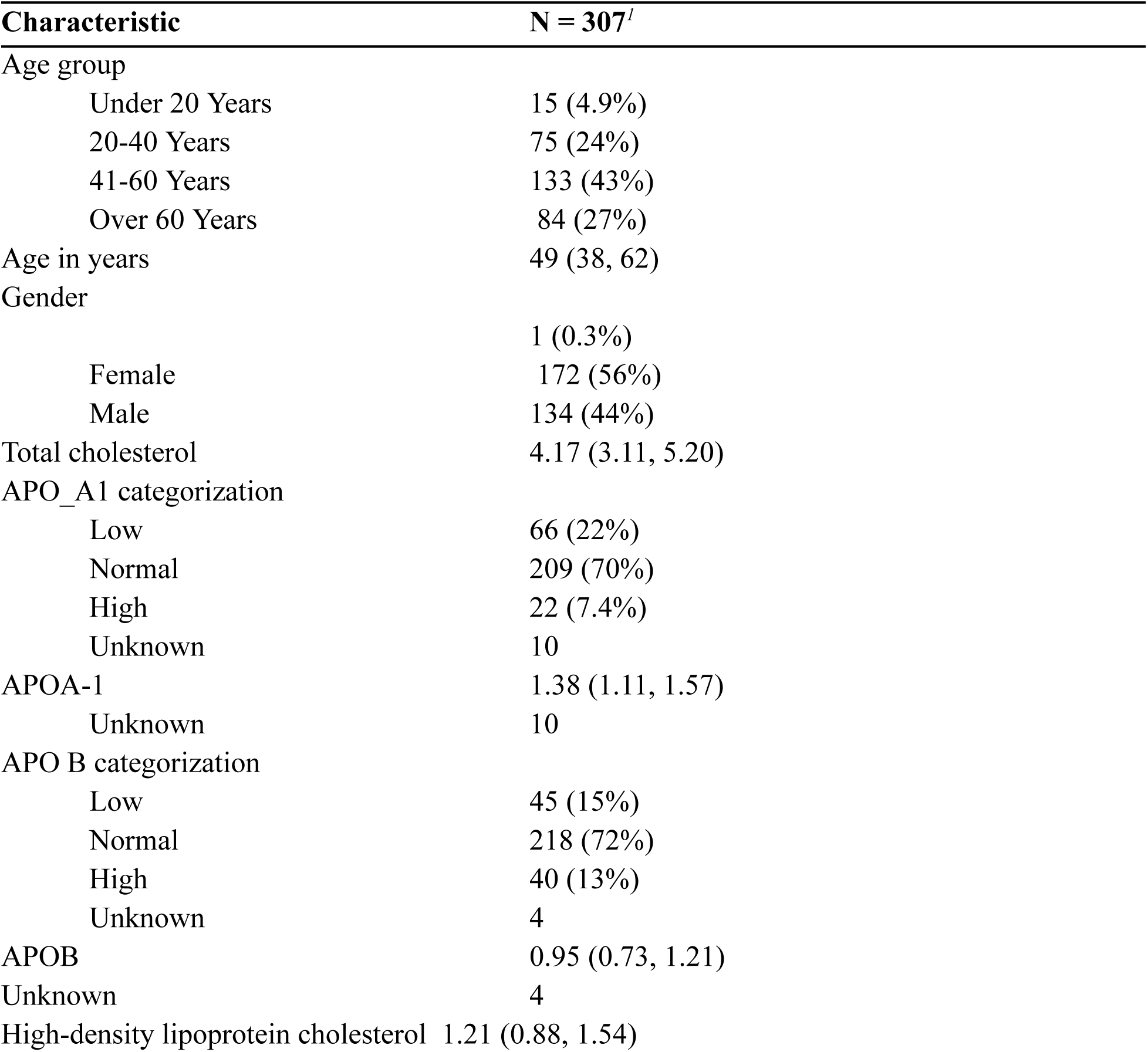
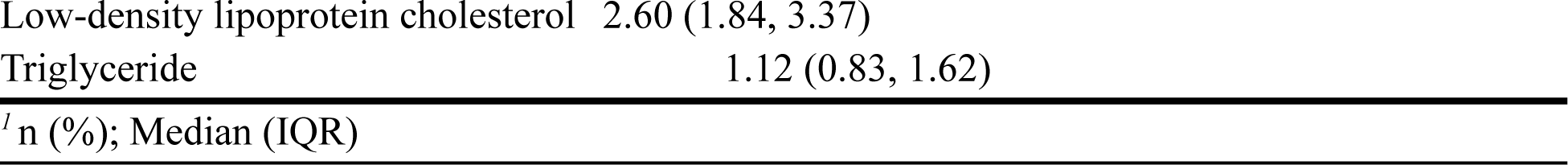
Baseline characteristics.

### Apo B / ApoA-1 ratio and atherogenic index in samples of patients sent routine lipid profile

The median value for APOB/APOA-1 ratio was 0.75 (0.56, 0.94). More than a third of the patients (42%, n=124) were at risk of CVD according to ApoB/ApoA-1 ratio. The atherogenic index had a median value of 0.09 (−0.14, 0.45). More than a third of the patients (40%, n=122) had a high CVD risk according to the atherogenic index stratification.

**Table 2:** Apo B/ApoA-1 ratio and atherogenic index in samples of patients sent routine lipid Profile.

| Characteristic | N = 307<sup>1</sup> |
| --- | --- |
| Apo B/ApoA-1 ratio |  |
| At risk | 124 (42%) |
| No risk | 170 (58%) |
| Unknown | 13 |
| Apo B/ApoA-1 ratio | 0.75 (0.56, 0.94) |
| Unknown | 13 |
| Atherogenic index group |  |
| Low risk | 158 (51%) |
| Intermediate risk | 27 (8.8%) |
| High risk | 122 (40%) |
| Atherogenic index | 0.09 (-0.14, 0.45) |
| <sup>1</sup>n (%); Median (IQR) |  |

The Spearman’s rank correlation results showed that there is a positive correlation between LDL-C and ApoA-1, Apo B and Apo B/ApoA-1 ratio.

Comparison of ApoA-1 and Apo B to LDL-C

**Table 3:** Comparison of ApoA-1 and Apo B to LDL-C.

| LDL-C |
| --- |

|  | Spearman, r | p-value |
| --- | --- | --- |
| ApoA-1 | 0.42 | <0.001 |
| Apo B | 0.87 | <0.001 |
| Apo B/ApoA-1 ratio | 0.52 | <0.001 |

### A comparison of Apo/ApoA-1 ratio with atherogenic index using the weighted kappa statistic

The weighted kappa results showed that there was statistically significant agreement between atherogenic index and APOB/APOA-1 ratio in CVD risk identification, kW=0.36 (95% CI, 0.27-0.46), p<0.001. The strength of agreement was fair.

**Table 4:**
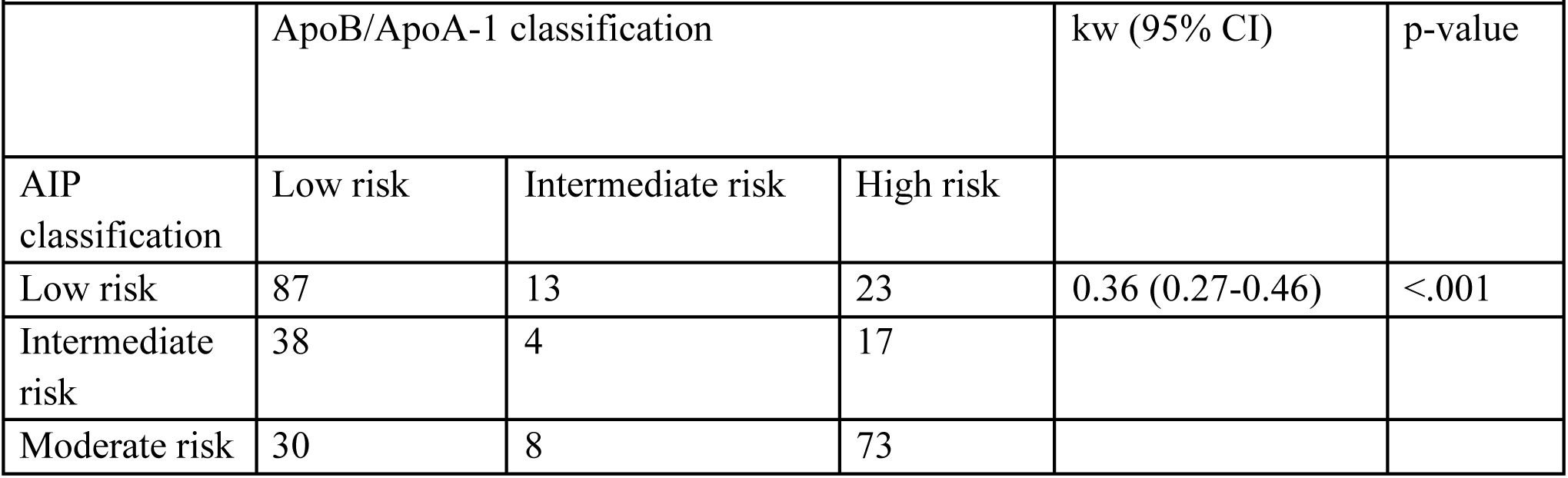
A comparison of ApoB /ApoA-1 ratio with atherogenic index using the weighted kappa statistic.

|  |  |  |  |  |  |
| --- | --- | --- | --- | --- | --- |
|  | ApoB/ApoA-1 classification |  |  | kw (95% CI) | p-value |
| AIP classification | Low risk | Intermediate risk | High risk |  |  |
| Low risk | 87 | 13 | 23 | 0.36 (0.27-0.46) | <.001 |
| Intermediate risk | 38 | 4 | 17 |  |  |
| Moderate risk | 30 | 8 | 73 |  |  |

### Risk for CHD

More than a quarter (26.7%) of the study participants had LDL-C values above the borderline high (2.35 mmol/l) for the classification of coronary heart disease risk.

**Table 5:**
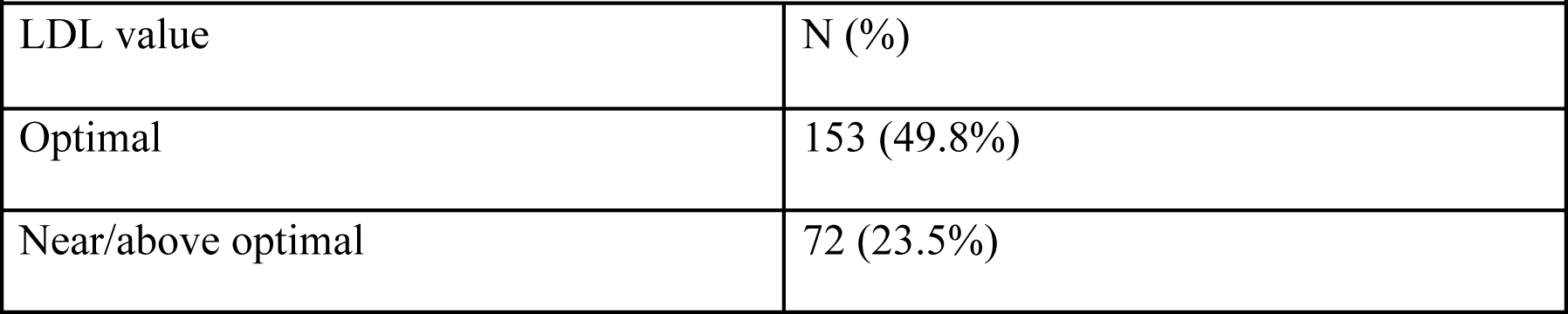
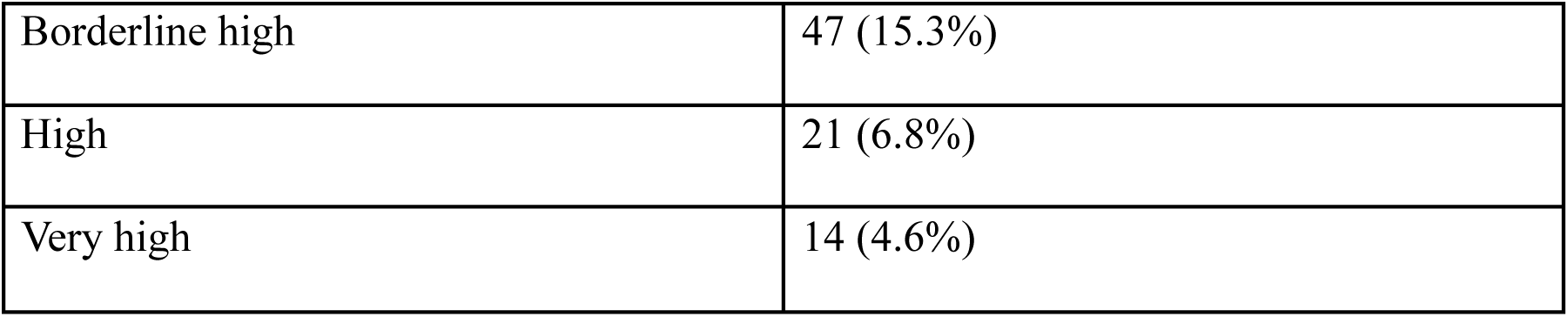
Risk of CHD by LDL-C classification.

| Table 5: Risk of CHD by LDL-C classification |  |
| --- | --- |
| LDL value | N (%) |
| Optimal | 153 (49.8%) |
| Near/above optimal | 72 (23.5%) |
| Borderline high | 47 (15.3%) |
| High | 21 (6.8%) |
| Very high | 14 (4.6%) |

## DISCUSSION

Globalization has led to a steady rise in cardiovascular diseases (CVD) in the low and middle income countries (LMICs). A number of risk factors are modifiable, dyslipidemia is one of them. The laboratory does analysis for a number of biomarkers that aid in the identification and monitoring of dyslipidemia in patients who are at risk of CVD. This current study sought to compare LDL-C, Atherogenic index of plasma (AIP) and Apo B/Apo A 1 ratio with an aim of establishing their role of identifying patients at risk of CVD in our population.

Our findings show that lipid profile is routinely requested at our biochemistry laboratory, the female gender (56%) had more requests than the male gender (44%) the median age of the study participants was 49 years with participants above 41 years accounting to 70% of the study population. This points out that lipid profile is routinely done in patients above the age of 40 years. Other studies (3, 19, 20, 21) also observed the similar age distribution in their study findings.

Examining the lipid profile parameter showed median (IQR) values as Total cholesterol 4.17 (3.11, 5.20), High-density lipoprotein cholesterol (HDL-C) 1.21 (0.88, 1.54) Low-density lipoprotein cholesterol (LDL-C) 2.60 (1.84, 3.37) Triglyceride 1.12 (0.83, 1.62). Assessment of these lipid profile values against our national guidelines (26) they do not indicate any risk for cardiovascular risk, though HDL-C values are borderline if they are to be examined against the female targets of the guideline. The distribution of LDL-C values for coronary heart disease risk were Optimal 153 (49.8%) Near/above optimal 72 (23.5%) Borderline high 47 (15.3%) High 21 (6.8%) Very high 14 (4.6%) this indicates that out of all the participants in the study 26.7% had borderline to very high LDL-C values.

Anastasiya M *et al* (22) explored this phenomenon and observed that patients with atherosclerotic conditions had their lipid profile within the respective population reference intervals. These lipid profile parameters are similar to findings by Niroumand S *et al* (3) who observed comparable values in an Iranian population except for LDL-C where their study observed slightly higher values.

The median Apo A was 1.38(1.11, 1.57) with normal levels being the majority (70%) and 22% being low and 7.4 %were found to be having high levels of Apo A using the reference interval of (1.05-2.05) .Apo B had a median of 0.95(0.73, 1.21) 455% had low levels, 72% were having normal levels while 13% were classified as having high levels using the reference interval of (0.55-1.30).It is key to note that the reference used for this interpretation were adopted from the assay reagent kit manufacturer.

Our study findings show that the median Apo B/Apo A ratio was 0.75(0.56, 0.96) Anastasiya M et al (36) found lower overall lower Apo B/Apo A-I values 0.52 (0.42; 0.78) compared to our findings in their study population. Using a cut off value of 0.8(19) the results showed that 42% of the study participants were classified as having a high risk of CVD. Other studies (19, 23, 24, and 25) have also shown similar findings underpinning the superiority Apo B/Apo A ratio in the CVD risk stratification in a diverse study population.

We also computed the atherogenic index of plasma (AIP) and the median value was 0.09 (−0.14, 0.45) and on using a cut off value of 0.21(40) 40% of the participants had a high CVD risk. Our findings are in contrast with Niroumand S *et al* (3) 77.5% of their study population having increased risk of CVD probably this could be due to the difference in the population studied.

Two other studies investigating AIP (44, 45) also observed differing results in their study done in a similar population as our current study found lower high risk participants of 22% and 25% respectively. This difference might be explained by the population where the study participants were drawn.

LDL-C showed a positive correlation with Apo A, Apo B and Apo A/Apo B ratio with the strongest correlation with Apo B (Spearman r of 0.87). This is explained by the fact that 90% or more of Apo B in circulation are found in low density lipoprotein (22)

We also compared the atherogenic index of plasma (AIP) and Apo B/Apo A ratio for cardiovascular disease risk identification and there was a fair and statistically significant agreement between the two biomarkers. This concurs Anastasiya M *et al* (22) who pointed out that AIP is the qualitative aspect of lipoproteins, while the ApoB/Apo A-I ratio is the quantitative part. Thus the agreement between the two (Apo B/Apo A-I ratio and AIP) indicates a direct relationship.

## STUDY LIMITATIONS

This was a cross sectional study on a general population drawn from patients presenting for lipid profile we did not classify the study population according to the underlying pathology or co morbidities and or ongoing therapy.

## CONCLUSION

Based on our study findings show that a standard lipid profile testing is inferior in the identification of patients at risk of cardiovascular disease. Apo A and Apo B markers used independently or in addition to the standard lipid profile can offer additional benefit in the CVD risk identification.

Atherogenic index of plasma (AIP) and Apo B/Apo A can be used to estimate and quantify atherogenic lipoproteins in plasma respectively.

## Data Availability

All relevant data are within the manuscript and its Supporting Information files.

